# Transcatheter Bicaval Valve Implantation For Treatment Of Severe Tricuspid Regurgitation: A Single Centre Registry

**DOI:** 10.64898/2026.05.26.26354174

**Authors:** Azmee B. Mohd Ghazi, Ow Ji Ken, Quah Wy Jin, Shaiful Azmi Yahaya

## Abstract

**Background:** Heterotopic caval valve implantation using the TricValve^®^ (OrbusNeich P&F) is a unique interventional approach for treatment of severe Tricuspid Regurgitation in patients who are deemed ineligible for surgery. Given the complexity and novelty of TricValve^®^ implantation, there is a pressing need for robust clinical data to evaluate its safety, efficacy, and long-term outcomes. Our study assesses the clinical results of patients followed up for 1 year from our centre.

**Methods:** Retrospective, single centre registry involving patients who have undergone TricValve^®^ Transcatheter Bicaval Valves System (OrbusNeich P&F) implantation for the treatment of severe tricuspid regurgitation.

**Results:** Fourteen patients were included. The mean age was 67.5 ± 8.7 years, with high surgical risk (mean EuroSCORE II 6.1 ± 3.7). Procedural success was achieved in thirteen patients, with no reported in-hospital mortality or stroke among all fourteen patients. At 1-year, significant improvements were observed in New York Heart Association (NYHA) functional class (86% Class III at baseline to 0% Class III at 1 year, P=0.002) and Kansas City Cardiomyopathy Questionnaire (KCCQ-12) scores (mean 32.0 ± 7.4 to 42.4 ± 12.0, P=0.015). TR Regurgitant Volume significantly decreased (65.5 ± 16.9 ml to 38.2 ± 13.6 ml, P=0.005). No deaths or strokes occurred during follow-up. Rehospitalization due to heart failure occurred in 14% (2 out of 14) of patients.

**Conclusion:** In this single-centre registry of high-risk patients, TricValve^®^ implantation was associated with a favorable safety profile, significant reduction in tricuspid regurgitant volume, and meaningful improvements in functional status and quality of life at 1 year.

**Clinical Perspective:** *What is New?:*

- This registry provides the first single-centre experience from a national heart centre in Asia evaluating the feasibility and safety of the TricValve transcatheter system.
- The study offers unique real-world data on procedural outcomes and patient selection specifically within an underrepresented Asia-Pacific population.
- Our findings demonstrate that caval valve implantation is a technically viable therapeutic option for Asian patients with severe tricuspid regurgitation and prohibitive surgical risk.

*What are the Clinical Implications?:*

- These results support the broader adoption of transcatheter caval therapies as a safe alternative for managing refractory right heart failure in the region.
- The observed short-term clinical improvements suggest that this intervention can effectively reduce the burden of recurrent hospitalizations in high-risk patients.
- This evidence contributes to the development of standardized procedural protocols for emerging right-sided structural heart programs in Asian medical centres.

## Introduction

Tricuspid regurgitation (TR) remains largely untreated due to clinical and anatomic limitations, although highly prevalent and has a significant impact on morbidity and mortality.^1,2^ The relatively higher risk of morbidity and mortality limits conventional surgery. ^1,3,4,5^ Patients are often ineligible for surgery due to age, previous cardiac surgery, late referral, comorbidities, pulmonary hypertension, and right ventricular (RV) dysfunction. Therefore, alternative approaches with transcatheter valve procedures have been increasingly applied in clinical practice.^6,7.^

Currently, isolated TR has emerged as a distinct entity, characterized by TR without other valvular heart disease.^8^ The increasing incidence of isolated tricuspid regurgitation (TR) has been attributed to the rising prevalence of atrial fibrillation (AF), intracardiac devices, and intravenous drug use. ^8,9,10,11,12,13,14^

To date, the two main approaches for percutaneous TR treatment are repair (leaflet approximation or annuloplasty) and replacement (orthotopic or heterotopic).^15^ Heterotopic caval valve implantation using the TricValve^®^ (OrbusNeich P&F) is a unique interventional approach for treatment of severe TR in patients who are deemed ineligible for surgery. The concept behind this approach is to reduce caval backflow, hence reducing systemic venous congestion promoting right ventricular remodelling and increasing cardiac output.^16^

Given the complexity and novelty of TricValve^®^ implantation, there is a pressing need for robust clinical data to evaluate its safety, efficacy, and long-term outcomes. Analysing real world data will provide insights on clinical practice, guide patient selection, and refine procedural techniques. We report on the retrospective clinical results of patients followed up for one year from our centre.

## Methods

### Study Design and Population

This study is designed as a retrospective, single centre registry involving patients with severe tricuspid regurgitation treated at National Heart Centre from 2021 to 2024, with follow-up period of one year post implantation. The inclusion criteria for the implantation were as follows: adult subjects with symptomatic (symptoms and signs of right heart failure and New York Heart Association [NYHA] functional class III or IV) severe or worse TR despite optimal medical therapy, suitable anatomic criteria for implantation assessed by computed tomography, Left Ventricular Ejection Fraction (LVEF) ≥ 35% assessed by echocardiography and patients’ ineligibility for other treatments after Heart Team discussion.

The exclusion criteria for the implantation were as follows: severe right ventricular dysfunction (tricuspid annular plane systolic excursion [TAPSE] ≤ 13 mm), a systolic pulmonary artery pressure > 65 mm Hg, known significant intracardiac shunt or congenital structural heart disease, need for other cardiac procedures (90 days after the procedure or 30 days before the procedure), patient life expectancy of less than one year, Cerebro-vascular event within the last three months, thrombocytopenia (absolute platelet count < 100,000/mm^3^) and patients with gastrointestinal bleeding within 90 days prior to screening.

The follow-up period extends to one-year post implantation to ensure a comprehensive evaluation of both short-term and long-term outcomes.

### Study Endpoints

The primary endpoint is intraprocedural success and was evaluated by the following: 1) absence of intraprocedural mortality or stroke; and 2) successful access, delivery, and retrieval of the device delivery system; and 3) successful deployment and correct positioning of the intended device(s) without requiring implantation of unplanned additional devices; and 4) adequate performance of the transcatheter device. Performance of devices whose purpose is a reduction in TR, should include the absence of tricuspid stenosis (TVA ≥ 1.5 cm^2^ or TVAi ≥ 0.9 cm2/m^2^ [≥ 0.75 if BMI >30 kg/m^2^], DVI <2.2, mean gradient <5 mm Hg); reduction of total tricuspid regurgitation to optimal (≤ mild [1+]) or acceptable (≤ moderate [2+]); 5) absence of device-related obstruction of forward flow; 6) Absence of device-related pulmonary embolism and 7) freedom from emergency surgery or reintervention during the first 24 h related to the device or access procedure.

The secondary endpoints are as follows: 1) Improvement from baseline in symptoms (e.g., NYHA improvement by ≥ 1 functional class); and/or Improvement from baseline in quality-of-life (e.g., Kansas City Cardiomyopathy Questionnaire improvement by ≥ 5); 2) freedom from major adverse events including death, acute myocardial infarction, tricuspid valve surgery, cardiac tamponade, stroke, or major bleeding according to Valve Academic Research Consortium-3 criteria at 1-year follow-up; and 3) freedom from heart failure rehospitalizations or serious adverse events related to the device at 1-year follow-up.

All clinical events were defined according to Valve Academic Research Consortium-3 criteria.

### Statistics

Baseline demographics, clinical characteristics, and procedural details were summarized using means and standard deviations for continuous variables, and frequencies and percentages for categorical variables. Subgroup analyses were performed on key demographics (e.g., age, gender) and clinical characteristics (e.g., comorbidities) to explore potential differences at baseline.

Paired t-tests were used to compare continuous variables (e.g., echocardiographic parameters) before and after implantation, Wilcoxon Signed-Rank Test were applied for non-normally distributed continuous data. McNemar’s Test was used to compare categorical outcomes (e.g., NYHA functional class) before and after implantation.

All statistical tests used a 2-sided alpha level of 0.05 as significance threshold for testing. Statistical analysis was performed using IBM SPSS software version 29.0.

### Procedure

After the patient was anesthetized, low doses of noradrenaline and milrinone were administered as inotropic support along with nitric oxide. Transoesophageal echocardiography (TEE) was performed to confirm previous findings. Bilateral transfemoral venous access was established, consisting of two punctures in the left femoral vein and one puncture in the right femoral vein. A Proglide closure device was deployed at the right femoral vein access site using a preclose technique in preparation for large-bore access. A multipurpose (MP) catheter was advanced via one of the left femoral vein accesses into the right pulmonary artery (rPA) to mark the crossing of the rPA with the superior vena cava (SVC). Through the second left femoral access, a 6 Fr pigtail catheter was advanced into the SVC, and a venogram was performed to delineate the venous anatomy and confirm appropriate valve landing zones. Via the right femoral venous access, a 0.035” soft wire was used to guide the MP catheter into the right internal jugular vein (RIJ). The soft wire was then exchanged for a Lunderquist extra-stiff wire (Cook Medical), after which the MP catheter was removed, leaving the Lunderquist wire in place. Over the stiff wire, sequential dilation of the right femoral vein was performed up to 22 Fr to accommodate the delivery system. The TricValve delivery system was introduced, and the SVC bioprosthesis was advanced and positioned at the SVC-right atrial junction, with the belly of the prosthesis placed above the rPA crossing. Final positioning was confirmed under fluoroscopic and transoesophageal echocardiographic (TOE/TEE) guidance, and the valve was then slowly deployed.

The delivery system was carefully withdrawn, and the MP catheter was also removed from the left femoral vein. Next the IVC bioprosthesis was loaded onto the delivery system and advanced through the right femoral venous access. The valve was positioned at the level of the diaphragm, with the prosthetic skirt visible just above the hepatic vein. Positioning was verified using TEE in the bicaval view, and the valve was successfully deployed. Final positioning confirmed that the prosthesis skirt extended approximately 20 mm into the right atrium, which is within acceptable limits for secure anchoring and function. Following successful deployment, haemostasis was achieved at the right femoral access site using the pre-deployed Proglide closure device.

## Results

A total of 14 patients with severe symptomatic tricuspid regurgitation (TR) who were considered high-risk for surgery were enrolled and underwent transcatheter bicaval valve implantation with the TricValve system. Baseline characteristics, procedural outcomes, changes in functional status, quality of life, laboratory parameters, echocardiographic findings, and adverse events at follow-up were assessed.

### Baseline Characteristics

The baseline demographic and clinical characteristics of the study cohort (N=14) are detailed in Table 1. The mean age was 67.5 ± 8.7 years, with an equal distribution of male and female patients. The mean BMI was 24.2 ± 3.9 kg/m2. Patients presented with high surgical risk profiles, indicated by a mean EuroSCORE II of 6.1 ± 3.7, STS score of 7.2 ± 5.2, and Tri-score of 6.1±1.8. The predominant baseline cardiac rhythm was permanent atrial fibrillation (64%), followed by sinus rhythm (29%) and paroxysmal atrial fibrillation (7%). The aetiology of TR was primarily ventricular (79%), with atrial, lead-related, and primary aetiologies each present in 7% of the cohort. Based on a 5-grade TR severity scale, baseline TR severity was categorized as Grade 3 in 21% of patients, Grade 4 in 43%, and Grade 5 in 36%.

**Table 1.** Baseline Characteristics (N=14)

|  |  |  |
| --- | --- | --- |
| 477 | Clinical |  |
| 478 | Age (years) | 67.5 ± 8.7 |
| 479 | Male/female | 7/7 |
| 480 | BMI | 24.2 ± 3.9 |
| 481 | EuroSCORE II | 6.1 ± 3.7 |
| 482 | STS score | 7.2 ± 5.2 |
| 483 | Tri-score | 6.1 ± 1.8 |
| 484 | Rhythm baseline |  |
| 485 | Sinus | 4 (29) |
| 486 | Paroxysmal Afib | 1 (7) |
| 487 | Permanent Afib | 9 (64) |
| 488 | TR aetiology |  |
| 489 | Atrial | 1 (7) |
| 490 | Ventricular | 11 (79) |
| 491 | Lead Related | 1 (7) |
| 492 | Primary | 1 (7) |
| 493 | TR Grade |  |
| 494 | Grade 3 | 3 (21) |
| 495 | Grade 4 | 6 (43) |
| 496 | Grade 5 | 5 (36) |
| 497 | Medical History |  |
| 498 | Hypertension | 14 (100) |
| 499 | Diabetes mellitus | 8 (57) |
| 500 | Chronic Kidney Disease | 11 (79) |
| 501 | Dialysis | 13 (93) |
| 502 | COPD | 0 (0) |
| 503 | Peripheral artery disease | 0 (0) |
| 504 | Coronary artery disease | 10 (71) |
| 505 | Stroke | 1 (7) |
| 506 | Pacemaker | 3 (21) |
| 507 | Prior hospitalization for heart failure | 10 (71) |
| 508 | Prior myocardial infarction | 4 (29) |
| 509 | Prior PCI | 3 (21) |
| 510 | Prior cardiac surgery | 6 (43) |
| 511 | Prior CABG | 3 (21) |
| 512 | Valve surgery |  |
| 513 | Aortic | 1 (7) |
| 514 | Mitral | 3 (21) |
| 515 | Tricuspid | 1 (7) |
| 516 | Percutaneous valve therapy |  |
| 517 | Aortic | 0 (0) |
| 518 | Mitral | 3 (21) |
| 519 | Tricuspid | 2 (14) |
| 520 | Values are mean ± SD, n (%), or median (Q1-Q3). AFib = atrial fibrillation; BMI = | |
| 521 | body mass index; CABG = Coronary artery bypass grafting; COPD = chronic |  |
| 522 | obstructive pulmonary disease; EuroSCORE II = European System for Cardiac |  |
| 523 | Operative Risk Evaluation II; PCI = Percutaneous Coronary Intervention; |  |

TRISCORE = Risk score model for isolated tricuspid valve surgery.

The cohort exhibited a high burden of comorbidities, including hypertension (100%), coronary artery disease (71%), diabetes mellitus (57%), and chronic kidney disease (79%), with most patients (93%) receiving dialysis at baseline. Prior cardiac history was common, with 71% having a history of prior hospitalization for heart failure, 29% with prior myocardial infarction, 21% with prior PCI, and 43% with prior cardiac surgery (including CABG in 21%). Previous valve interventions included mitral valve surgery (21%), tricuspid valve surgery (7%), percutaneous mitral valve therapy (21%), and percutaneous tricuspid valve therapy (14%).

### Functional Class, Quality of Life and Laboratory Parameters

Changes in NYHA functional class and quality of life (measured by the KCCQ-12 score) are presented in Table 2 and Figure 1. A statistically significant improvement in NYHA functional class was observed from baseline to the first follow-up (P=0.002) and was maintained through the last follow-up (P=0.002). At baseline, 86% of patients were in NYHA Class III. This proportion decreased markedly at the first follow-up, with 77% in Class II and 15% in Class I. At the last follow-up, 70% were in Class II and 30% were in Class I, with no patients remaining in Class III.

**Figure 1:**
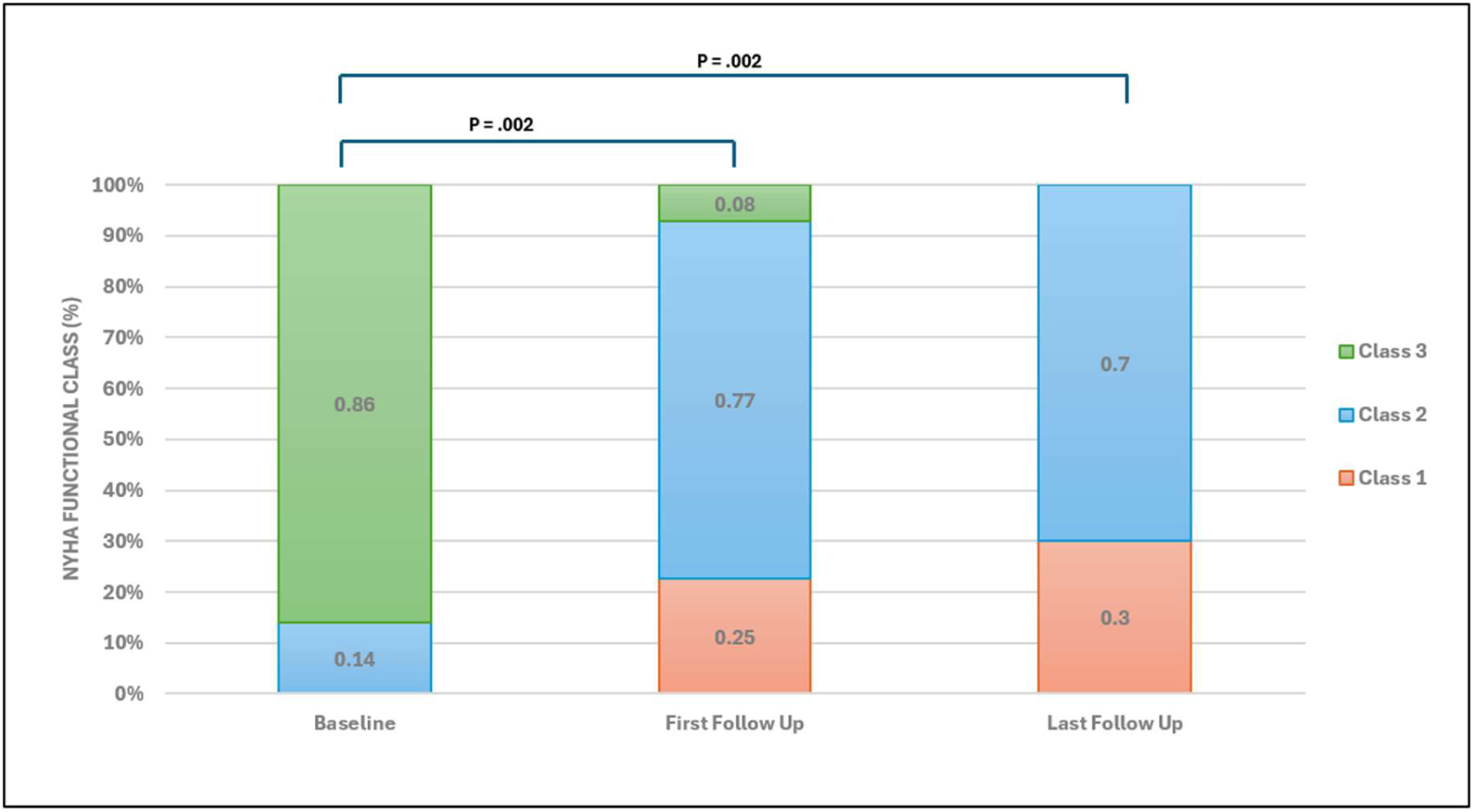
NYHA Functional Class at Baseline and During Follow Up. Legend: Baseline, first follow up, and last follow-up in NYHA functional class. Data are presented as %.

**Table 2.** Functional Class, Quality of Life and Laboratory Parameters.

|  | Baseline | First Follow Up | p-value | Last Follow Up | p-value |
| --- | --- | --- | --- | --- | --- |
| Functional Class |  |  |  |  |  |
| NYHA Class I | 0 (0) | 2 (15) |  | 3 (30) |  |
| NYHA Class II | 2 (14) | 10 (77) | 0.002 | 7 (70) | 0.002 |
| NYHA Class III | 12 (86) | 1 (8) |  | 0 (0) |  |
| Quality of Life |  |  |  |  |  |
| KCCQ-12 score | 32.0 ± 7.4 | 38.1 ± 7.0 | 0.038 | 42.4 ± 12.0 | 0.015 |
|  | 30.5 (27.3 - 40.0) | 36 (32.0 - 44.5) | 0.044 | 37.5 (32.5 - 55.0) | 0.020 |
| Laboratory Parameters |  |  |  |  |  |
| eGFR, ml/min | 43.0 ± 15.5 | 47.7 ± 15.3 | 0.429 | 46.5 ± 8.1 | 0.535 |
| ALT, U/L | 19.1 ± 9.6 | 16.1 ± 7.8 | 0.382 | 15.3 ± 4.8 | 0.265 |
| NT-ProBNP, pg/mL | 3714 (1764 - 8347) | 2907 (1484 - 4319) | 0.381 | 1747 (772 - 2947) | 0.070 |
| Creatinine, mg/dL | 1.6 ± 1.0 | 1.6 ± 1.3 | 0.929 | 1.4 ± 0.4 | 0.389 |

Values are mean ± SD, n (%), or median (Q1-Q3).
ALT = alanine aminotransferase; eGFR = glomerular filtration rate; KCCQ-12 = 12-item Kansas
City Cardiomyopathy Questionnaire; NT-proBNP = N-terminal pro-B-type natriuretic peptide.

Figure 1 graphically displays the shift in NYHA functional class distribution over the course of the study, highlighting the reduction in patients in higher functional classes and the increase in those in lower classes at follow-up. Quality of life, assessed using the KCCQ-12 score, demonstrated significant improvement from baseline to both follow-up periods (Table 2). The mean KCCQ-12 score increased from 32.0 ± 7.4 at baseline to 38.1 ± 7.0 at the first follow-up (P=0.038) and to 42.4±12.0 at the last follow-up (P=0.015). Similarly, the median KCCQ-12 score increased from 30.5 (IQR 27.3-40.0) at baseline to 36 (IQR 32.0-44.5) at the first follow-up (P=0.044) and 37.5 (IQR 32.5-55.0) at the last follow-up (P=0.020).

Changes in key laboratory parameters are also shown in Table 2. No statistically significant changes were observed in mean eGFR, ALT, NT-ProBNP, or Creatinine levels from baseline to either the first or last follow-up. The median NT-ProBNP showed a trend towards reduction at the last follow-up (1747 pg/mL vs 3714 pg/mL at baseline), although this did not reach statistical significance (P=0.070).

### Echocardiographic Parameters

Table 3 provides a detailed assessment of echocardiographic parameters at baseline and the last follow-up. Regarding tricuspid valve and regurgitation assessment, a statistically significant reduction was observed in TR regurgitant volume, decreasing from 65.5 ± 16.9 ml at baseline to 38.2±13.6 ml at the last follow-up (P=0.005). Other TR quantification parameters, including vena contracta (10.6 ± 3.5 mm at baseline vs 8.8 ± 2.2 mm at last follow-up, P=0.294) and effective regurgitant orifice area (EROA) (proximal isovelocity surface area [PISA], 98.5 ± 42.8 mm^2^ at baseline vs 81.5 ± 38.2 mm^2^ at last follow-up, P=0.437), showed numerical reductions but did not reach statistical significance. Tricuspid annulus diameter and tricuspid valve (TV) coaptation gap also did not show statistically significant changes.

**Table 3.**
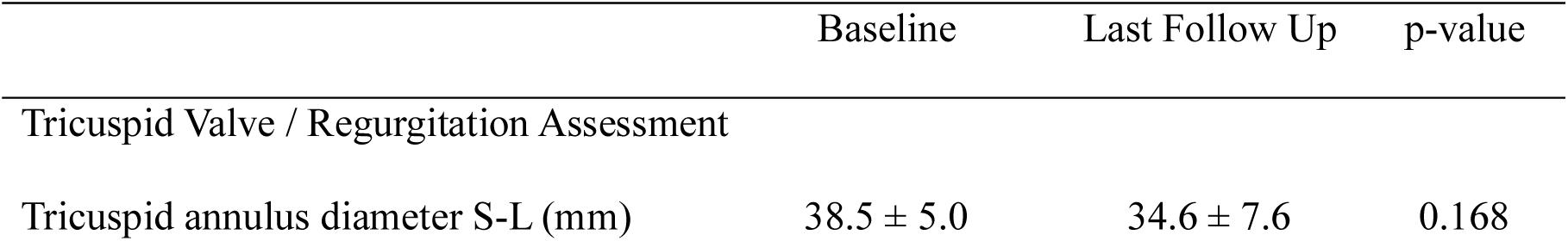
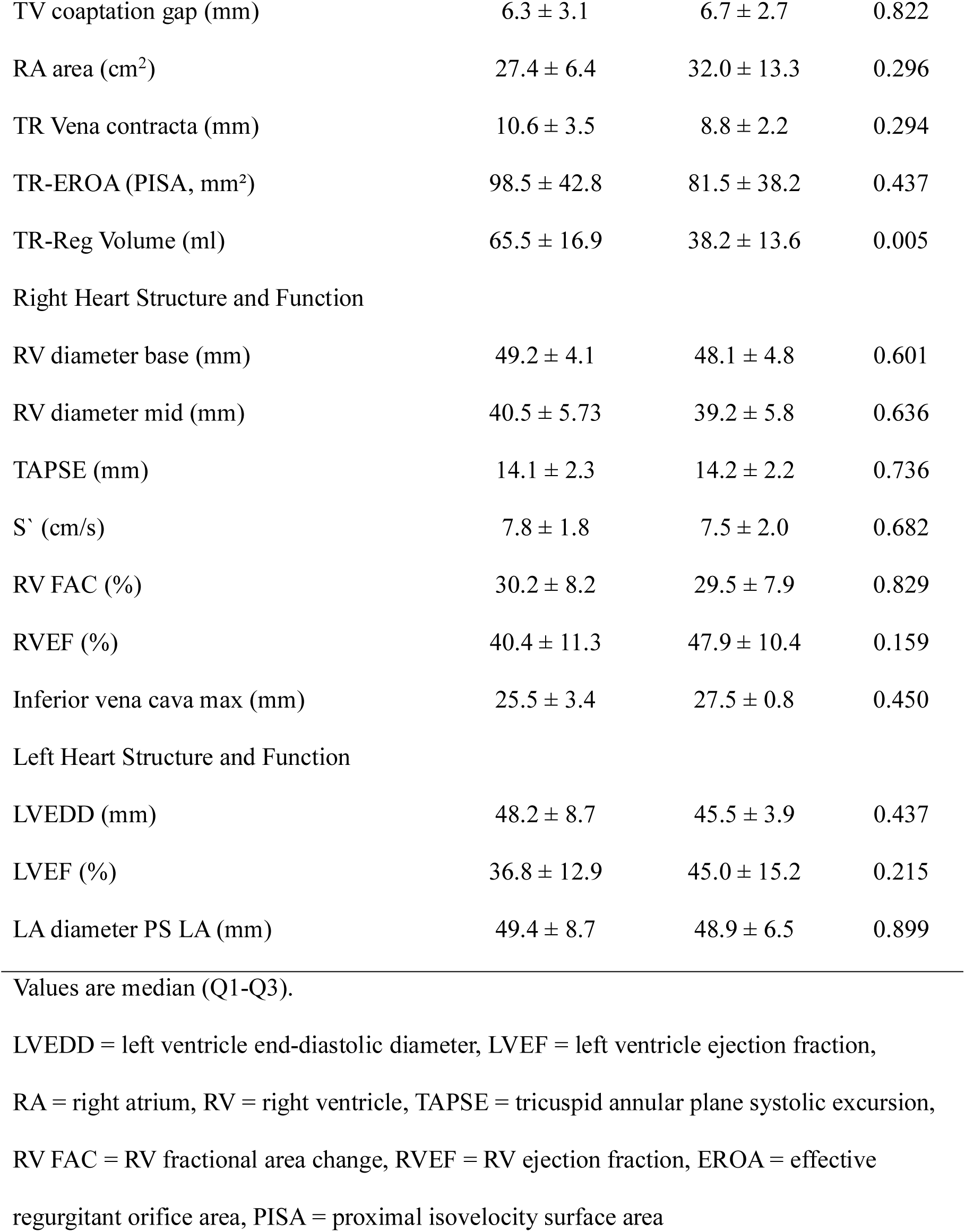
Echocardiographic Parameters: Baseline and Last Follow-Up.

|  | Baseline | Last Follow Up | p-value |
| --- | --- | --- | --- |
| Tricuspid Valve / Regurgitation Assessment |  |  |  |
| Tricuspid annulus diameter S-L (mm) | 38.5 ± 5.0 | 34.6 ± 7.6 | 0.168 |

|  |  |  |  |  |
| --- | --- | --- | --- | --- |
| 548 | TV coaptation gap (mm) | 6.3 ± 3.1 | 6.7 ± 2.7 | 0.822 |
| 549 | RA area (cm<sup>2</sup>) | 27.4 ± 6.4 | 32.0 ± 13.3 | 0.296 |
| 550 | TR Vena contracta (mm) | 10.6 ± 3.5 | 8.8 ± 2.2 | 0.294 |
| 551 | TR-EROA (PISA, mm<sup>2</sup>) | 98.5 ± 42.8 | 81.5 ± 38.2 | 0.437 |
| 552 | TR-Reg Volume (ml) | 65.5 ± 16.9 | 38.2 ± 13.6 | 0.005 |
| 553 | Right Heart Structure and Function |  |  |  |
| 554 | RV diameter base (mm) | 49.2 ± 4.1 | 48.1 ± 4.8 | 0.601 |
| 555 | RV diameter mid (mm) | 40.5 ± 5.73 | 39.2 ± 5.8 | 0.636 |
| 556 | TAPSE (mm) | 14.1 ± 2.3 | 14.2 ± 2.2 | 0.736 |
| 557 | S' (cm/s) | 7.8 ± 1.8 | 7.5 ± 2.0 | 0.682 |
| 558 | RV FAC (%) | 30.2 ± 8.2 | 29.5 ± 7.9 | 0.829 |
| 559 | RVEF (%) | 40.4 ± 11.3 | 47.9 ± 10.4 | 0.159 |
| 560 | Inferior vena cava max (mm) | 25.5 ± 3.4 | 27.5 ± 0.8 | 0.450 |
| 561 | Left Heart Structure and Function |  |  |  |
| 562 | LVEDD (mm) | 48.2 ± 8.7 | 45.5 ± 3.9 | 0.437 |
| 563 | LVEF (%) | 36.8 ± 12.9 | 45.0 ± 15.2 | 0.215 |
| 564 | LA diameter PS LA (mm) | 49.4 ± 8.7 | 48.9 ± 6.5 | 0.899 |

Values are median (Q1-Q3).
LVEDD = left ventricle end-diastolic diameter, LVEF = left ventricle ejection fraction, RA = right atrium, RV = right ventricle, TAPSE = tricuspid annular plane systolic excursion, RV FAC = RV fractional area change, RVEF = RV ejection fraction, EROA = effective regurgitant orifice area, PISA = proximal isovelocity surface area

Parameters assessing right heart structure and function, including right ventricle (RV) diameter at the base and mid-cavity, tricuspid annular plane systolic excursion (TAPSE), S’ velocity, RV fractional area change (FAC), and right ventricular ejection fraction (RVEF), did not demonstrate statistically significant changes from baseline to the last follow-up. The maximum diameter of the inferior vena cava numerically increased from 25.5 ± 3.4 mm at baseline to 27.5 ± 0.8 mm at the last follow-up, but this was not statistically significant (P=0.450). Right atrial area increased from 27.4 ± 6.4 cm^2^ at baseline to 32.0 ± 13.3 cm^2^ at the last follow-up, but this change was not statistically significant (P=0.296). Left heart structure and function parameters similarly did not show significant changes during the study period.

### Procedural Outcomes

As shown in Table 4, intraprocedural success was achieved in 13 out of 14 patients (92.9%). There was one patient with access complications (right inguinal hematoma, with small pseudoaneurysm) during the procedure. There was no reported in-hospital mortality, stroke/TIA, cardiac complications, new pacemaker implantation, inadequate performance of the transcatheter device, device-related obstruction of forward flow, pulmonary embolization, or conversion to surgery or reintervention. All patients remained under anticoagulant treatment at the time of discharge.

**Table 4.** Intraprocedural Parameters.

|  |  |  |
| --- | --- | --- |
| 572 | In-hospital mortality | 0 (0) |
| 573 | Stroke/TIA | 0 (0) |
| 574 | Access complications | 1 (7) |
| 575 | Cardiac complications | 0 (0) |
| 576 | New pacemaker implantation | 0 (0) |
| 577 | Device embolization/migration | 0 (0) |
| 578 | Inadequate performance of the transcatheter device | 0 (0) |
| 579 | Device-related obstruction of forward flow | 0 (0) |
| 580 | Pulmonary embolization | 0 (0) |
| 581 | Conversion to surgery or reintervention | 0 (0) |

Values are n (%).
TVARC defines intraprocedural success as successful deployment and adequate immediate performance of the device in the absence of serious complications, and clinical success as the proper positioning of the device with adequate device function and the absence of procedure-related complications, need for reintervention, or readmissions for the underlying condition.

### Major and Serious Adverse Events

Table 5 summarizes the major and serious adverse events reported during the follow-up period. There were no occurrences of major adverse events such as all-cause death, myocardial infarction, tricuspid valve surgery, cardiac tamponade, or stroke at any point during the follow-up. One case of major bleeding occurred procedurally (7% overall 1 year), and one instance of shoulder pain was noted procedurally (7% overall 1 year). As for serious adverse events, heart failure rehospitalization occurred in one patient at the first follow-up and another at the last follow-up, resulting in an overall 1-year rate of 14%. One other serious adverse event was reported at the first follow-up (7% overall 1 year). There were no reported paravalvular leaks.

**Table 5.** Major and Serious Adverse Events.

| 591 |  | Procedural | First | Last | Total |
| --- | --- | --- | --- | --- | --- |
| 592 |  |  | Follow Up | Follow Up | 1 year (%) |
| 593 | Major Adverse Event |  |  |  |  |
| 594 | All-cause death | 0 | 0 | 0 | 0% |
| 595 | Myocardial Infarction | 0 | 0 | 0 | 0% |
| 596 | Tricuspid valve surgery | 0 | 0 | 0 | 0% |
| 597 | Cardiac tamponade | 0 | 0 | 0 | 0% |
| 598 | Major bleeding | 1 | 0 | 0 | 7% |
| 599 | Stroke | 0 | 0 | 0 | 0% |
| 600 | Serious Adverse Event |  |  |  |  |
| 601 | Heart failure rehospitalization | 0 | 1 | 1 | 14% |
| 602 | Shoulder pain | 1 | 0 | 0 | 7% |
| 603 | Paravalvular leak | 0 | 0 | 0 | 0% |
| 604 | Other procedural/in-hospital events | 0 | 1 | 0 | 7% |

## Discussion

This study presents the results of transcatheter bicaval valve implantation with the TricValve system in a cohort of 14 high-risk patients with severe symptomatic tricuspid regurgitation. The findings demonstrate favourable procedural safety and significant improvements in functional status and quality of life at follow-up, aligning with the goals of therapy for this challenging patient population as outlined by the TVARC definitions.

The patient cohort in this study represents a group with significant comorbidities and high surgical risk, consistent with the intended patient population for transcatheter TR interventions. The high prevalence of conditions such as chronic kidney disease requiring dialysis, prior cardiac surgery, and atrial fibrillation underscores the medical fragility of these patients, who are often deemed unsuitable for conventional surgical TV intervention.

The procedural outcomes observed in this study, with a low incidence of major adverse events, are comparable to the favourable safety profiles reported in larger studies of the TricValve system. The TRICUS EURO study (N=35) reported a procedural success rate of 94% with no procedural deaths or conversions to surgery.^19^ The combined TRICUS and TRICUS EURO 1-year follow-up analysis (N=44) similarly noted a low all-cause mortality rate (6.8%) and heart failure rehospitalization rate (29.5%) at 1 year.^18,19^ While the sample size in our study is smaller, the absence of in-hospital or follow-up mortality and stroke is encouraging. The observed access complications rate (7%) highlights potential procedural challenges that may decrease with increasing operator experience, as suggested by the learning curve discussed in the TRICUS/TRICUS EURO studies. ^18,19^

A key finding of this study is the significant and sustained improvement in NYHA functional class and KCCQ-12 scores. This aligns with the primary endpoints and results of the TRICUS EURO study, which demonstrated significant increases in KCCQ scores and improvement in NYHA class at 6 months. ^18,19^ The combined 1-year TRICUS/TRICUS EURO analysis further supports these findings, with 95.5% of patients achieving clinical improvement based on a composite endpoint including KCCQ score and NYHA class. The observed improvements in functional status and quality of life are clinically meaningful and address key patient-centred outcomes relevant to the evaluation of TR therapies. The TVARC document emphasizes the importance of patient-reported outcomes like KCCQ in assessing the effectiveness of new devices, even noting that improvement in health status alone can demonstrate effectiveness.

Echocardiographic assessment in our study revealed a statistically significant reduction in TR regurgitant volume at the last follow-up. While other TR quantification parameters like vena contracta and EROA showed numerical reductions that were not statistically significant, the reduction in regurgitant volume is a positive indicator of the intervention’s impact on TR severity. The TRICUS EURO study reported a lower percentage of at least severe TR at 6 months. ^19^ The combined 1-year analysis highlighted the abolition of hepatic vein backflow in a significant proportion of patients, contributing to reduced congestion. While our study did not specifically report on hepatic vein flow reversal, the reduction in TR volume is consistent with the mechanism of action of the TricValve in reducing caval backflow. The lack of significant changes in most right heart structural and functional parameters at the observed follow-up in our small cohort is consistent with findings in the TRICUS EURO study, where significant right heart remodelling was not observed at 6 months. The combined 1-year analysis also showed no significant changes in RV function by TAPSE, although a significant reduction in tricuspid annular diameter was noted. The TVARC document discusses the challenges in assessing RV size and function and the variable impact of TR interventions on right heart remodelling. Longer-term follow-up may be necessary to fully assess the effects on right heart reverse remodelling.

This study has several limitations, including the small sample size, which may limit the generalizability of the findings and the power to detect statistically significant changes in all parameters. The follow-up duration is also relatively short compared to the expected lifespan of the device and the chronic nature of severe TR. However, these results contribute to the growing body of evidence supporting the use of the TricValve system as a valuable treatment option for carefully selected high-risk patients with severe symptomatic TR who have limited alternative treatment options.

## Conclusion

In conclusion, the results of this study, in line with findings from larger trials like TRICUS EURO and the combined TRICUS/TRICUS EURO analysis, indicate that transcatheter bicaval valve implantation with the TricValve system is associated with a favourable safety profile and significant improvements in functional status and quality of life in a high-risk patient population with severe symptomatic TR. Future research with larger cohorts and longer follow-up periods, including ongoing registries and planned randomized trials, will further clarify the long-term durability of these benefits, the impact on clinical outcomes such as mortality and rehospitalization, and help define the optimal role of CAVI within the evolving treatment paradigm for severe TR. The adoption of standardized definitions and endpoints, as recommended by TVARC, will be crucial for meaningful comparisons across studies and advancing the understanding of this complex disease and its management.

## Future Research Directions

Future research should focus on several key areas to further elucidate the long-term efficacy and safety of the TricValve system. Larger, multicentre randomized controlled trials with extended follow-up periods are essential to confirm the findings of this study and provide more robust data on clinical outcomes such as mortality, rehospitalization, and quality of life. Additionally, studies should aim to identify patient subgroups that may derive the most benefit from this intervention, as well as those who may be at higher risk for adverse events.

Investigations into the mechanisms underlying the observed improvements in functional status and quality of life, including detailed hemodynamic assessments and imaging studies, will be valuable in understanding the physiological impact of the TricValve system. Furthermore, research into the potential for right heart reverse remodelling and the long-term durability of the device will be critical in establishing its role in the management of severe TR.

Finally, the development and validation of standardized definitions and endpoints, as recommended by TVARC, will be crucial for meaningful comparisons across studies and advancing the understanding of this complex disease and its management. Collaborative efforts among researchers, clinicians, and industry partners will be essential in driving innovation and improving outcomes for patients with severe symptomatic TR.

## Acknowledgements

We would like to acknowledge the cardiac anaesthesia/intensive team led by Dato’ Dr. Suhaini Kadiman, Dato’ Dr N. Thiru Kumar A/L A. Namasiwayam, and Dato’ Dr Suneta Sulaiman as well as the cardiothoracic team led by Prof. Dato’ Dr Mohamed Ezani Md Taib, Dato’ Dr Mohd Nazeri Nordin, and Prof Dato’ Sri Dr Jeffrey Jesswant Dillon for their support and help in making this endeavour successful. A special thanks to our dedicated senior echocardiographer Mr Deventhiran Permal. We would also like to extend our gratitude to the proctor Dr Ignacio J. Amat Santos from Spain for his guidance.

## Sources of Funding

The authors received no financial support for the research, authorship, and/or publication of this article.

## Disclosures

The authors have no conflicts of interest to declare that are relevant to the content of this article.

## Data availability statement

The data underlying this article will be shared on reasonable request to the corresponding author.

## Non-standard Abbreviation and Acronyms

CAVI: Caval Valve Implantation
EROA: Effective regurgitant orifice area
FAC: Fractional area change
MP: Multipurpose
PISA: Proximal isovelocity surface area
TAPSE: Tricuspid annular plane systolic excursion
TRISCORE: Risk score model for isolated tricuspid valve surgery
TTVR: Transcatheter Tricuspid Valve Replacement
TVARC: Tricuspid Valve Academic Research Consortium

## Notes

### Competing Interest Statement

The authors have declared no competing interest.

### Clinical Trial

The study was registered with local IRB and received approval.

### Author Declarations

Institut Jantung Negara Research Ethics Committee (IJNREC)

## References

1. Nath, J, Foster, E, Heidenreich, PA. Impact of tricuspid regurgitation on long-term survival. J Am Coll Cardiol. 2004;43:405–409. doi: 10.1016/j.jacc.2003.09.036.

2. Topilsky, Y, Maltais, S, Medina Inojosa, J, Oguz, D, Michelena, H, Maalouf, J, Mahoney, DW, Enriquez-Sarano, M. Burden of tricuspid regurgitation in patients diagnosed in the community setting. JACC Cardiovasc Imaging. 2019;12:433–442. doi: 10.1016/j.jcmg.2018.06.014.

3. Kwak JJ, Kim YJ, Kim MK, Kim HK, Park JS, Kim KH, Kim KB, Ahn H, Sohn DW, Oh BH, Park YB. Development of tricuspid regurgitation late after left-sided valve surgery: a single-centre experience with long-term echocardiographic examinations. Am Heart J 2008;155:732–7.

4. Bruce CJ, Conolly HM. Right-sided valve disease deserves a little more respect. Circulation 2009;119:2726–34.

5. Topilsky Y, Nkomo VT, Vatury O, Michelena HI, Letourneau T, Suri RM, Pislaru S, Park S, Mahoney DW, Biner S, Enriquez-Sarano M. Clinical outcome of isolated tricuspid regurgitation. JACC Cardiovasc Imaging 2014;7:1185–94.

6. Généreux P, Head SJ, Wood DA, Kodali SK, Williams MR, Paradis JM, Spaziano M, Kappetein AP, Webb JG, Cribier A, Leon MB. Transcatheter aortic valve implantation 10-year anniversary: review of current evidence and clinical implications. Eur Heart J 2012;33:2388–98.

7. Maisano F, La Canna G, Colombo A, Alfieri O. The evolution from surgery to percutaneous mitral valve interventions: the role of the edge-to-edge technique. J Am Coll Cardiol 2011;58:2174–82.

8. M. Di Mauro, G. Bonalumi, I. Giambuzzi, G. Masiero, G. Tarantini, Isolated tricuspid regurgitation: a new entity to face. Prevalence, prognosis and treatment of isolated tricuspid regurgitation, Minerva Cardiol. Angiol. (2023 Apr 6), 10.23736/S2724-5683.23.06294-4.

9. A.M. Calafiore, S. Gallina, A.L. Iac’o, et al., Mitral valve surgery for functional mitral regurgitation: should moderate-or-more tricuspid regurgitation be treated? A propensity score analysis, Ann. Thorac. Surg. 87 (2009) 698–703, 10.1016/j.athoracsur.2008.11.028.

10. J. Dreyfus, M. Flagiello, B. Bazire, et al., Isolated tricuspid valve surgery: impact of aetiology and clinical presentation on outcomes, Eur. Heart J. 41 (2020) 4304–4317, 10.1093/eurheartj/ehaa643.

11. M. Di Mauro, M. Russo, G. Saitto, et al., Prognostic role of endocarditis in isolated tricuspid valve surgery. A propensity-weighted study, Int. J. Cardiol. (2022), 10.1016/j.ijcard.2022.09.020.

12. D. Paniagua, H.R. Aldrich, E.H. Lieberman, G.A. Lamas, A.S. Agatston, Increased prevalence of significant tricuspid regurgitation in patients with transvenous pacemakers leads, Am. J. Cardiol. 82 (1130–2) (1998) A9, 10.1016/s0002-9149(98)00567-0.

13. Y. Topilsky, V.T. Nkomo, O. Vatury, et al., Clinical outcome of isolated tricuspid regurgitation, JACC Cardiovasc. Imaging 7 (2014) 1185–1194, 10.1016/j.jcmg.2014.07.018.

14. M. Russo, M. Di Mauro, G. Saitto, et al., Outcome of patients undergoing isolated tricuspid repair or replacement surgery, Eur. J. Cardiothorac. Surg. (2022) 62, 10.1093/ejcts/ezac230.

15. Curio J, Demir OM, Pagnesi M, Mangieri A, Giannini F, Weisz G, Latib A. Update on the Current Landscape of Transcatheter Options for Tricuspid Regurgitation Treatment. Interv Cardiol. 2019;14:54–61.

16. Lauten A, Ferrari M, Hekmat K, Pfeifer R, Dannberg G, Ragoschke-Schumm A, Figulla HR. Heterotopic transcatheter tricuspid valve implantation: first-in-man application of a novel approach to tricuspid regurgitation. Eur Heart J. 2011;32:1207–13.

17. P&F Products & Features. (2019, May 27). TricValve® Transcatheter Bicaval Valves System Overview. Siprotec.com.ar.

18. Estévez-Loureiro, R, Sánchez-Recalde, A, Amat-Santos, I. et al. 6-Month Outcomes of the TricValve System in Patients With Tricuspid Regurgitation: The TRICUS EURO Study. J Am Coll Cardiol Intv. 2022 Jul, 15 (13) 1366–1377.

19. Blasco-Turrión, S, Briedis, K, Estévez-Loureiro, R. et al. Bicaval TricValve Implantation in Patients With Severe Symptomatic Tricuspid Regurgitation: 1-Year Follow-Up Outcomes. J Am Coll Cardiol Intv. 2024 Jan, 17 (1) 60–72.

